# Benchmark Self-Triage (BeST): Comparing Self-Triage Systems and Multiple Generations of OpenAI Models Against Preliminary Expert Consensus

**DOI:** 10.64898/2026.10.12.26365287

**Authors:** Lars Eric Kroll, Anja Seidel, Josephine Schuster, Edgar Steiger, Timo Schöpke, Dominik von Stillfried, BeST Study Group

## Abstract

As digital self-triage becomes more widely used, quality assurance requires evaluation of both emergency recognition and overall agreement with clinical judgement, including how these outcomes differ across generations of large language models. The Benchmark Self-Triage (BeST) project intends to provide a framework to compare systems for digital self-triage against a medical ‘gold standard’ in Germany. As a first step in developing and demonstrating the methodology, we compared study-specific implementations of SmED 5.4.11 accessed via API and Infermedica Engine API via Microsoft Azure Healthcare Agent Service with five structured OpenAI configurations spanning multiple model generations: gpt-4.1, gpt-5.4, gpt-5.6-luna, gpt-5.6-sol, and gpt-6-astra. The manufacturers of SmED and Infermedica offer MDR-certified Class IIb triage products, but these certified products were not directly evaluated. An LLM-based patient actor covered 480 physician-curated vignettes derived from German routine triage data. A pool of seven physicians provided preliminary reference classifications and consensus for 480 vignettes. Recommendations were mapped to three urgency categories-emergency, acute, and not urgent-and evaluated using winsorised weights derived from routine triage frequencies. Weighted emergency sensitivity ranged from 51.3% (95% CI 35.566.3%) for gpt-4.1 to 99.9% (99.7-100.0%) for gpt-6-astra. Weighted exact three-category agreement (accuracy) ranged from 54.4% to 68.8%; gpt-4.1 achieved the highest agreement despite the lowest emergency sensitivity. No system performed best across all urgency categories. The later-generation OpenAI configurations showed a more precautionary triage profile resembling SmED, with higher sensitivity for emergency cases; gpt-6-astra showed the strongest shift. This cross-sectional comparison across model generations suggests a shift towards more precautionary triage behaviour, without establishing a longitudinal improvement in accuracy. These findings highlight the limitations of overall agreement for evaluating self-triage and support category-specific assessment. Broader expert consensus and evaluation, as well as further steps to reduce dependence on SmED-derived source data, are needed and planned before drawing conclusions about clinical safety.

## Introduction

Self-triage systems can influence whether people opt for emergency, urgent outpatient, or self-care pathways. Their evaluation therefore needs to assess the ability to identify both high-acuity presentations and cases that do not require urgent care, or no professional care at all. Earlier studies have found heterogeneity in symptom-checker triage accuracy^1,2^, and structured comparisons now include large language models (LLMs)^3,4^.

Like other European health systems based on social insurance Germany has established a central medical helpline to guide individuals seeking health advice or care to the appropriate service^5–7^. Software-based telephone triage, initially introduced to reduce avoidable utilisation of emergency care, is increasingly being supplemented by digital self-triage as a means to streamline access to care^8^. The Federal Government’s draft Act for Data and Digital Innovation in Healthcare (Gesetz für Daten und digitale Innovation im Gesundheitswesen; GeDIG) proposes a new Section 360b of Social Code Book V, under which the National Association of Statutory Health Insurance Physicians and the National Association of Statutory Health Insurance Funds are to agree requirements for a standardised, structured digital assessment of care needs^9^. This is intended to become a central entry point to the staturory health care system, comparable to the NHS app and the 111 helpline in England^10^. The proposed requirements explicitly include accuracy, including sensitivity and specificity, alongside patient safety, reliability and reproducibility, lay comprehensibility, explainability, and impartiality. Benchmark Self-Triage (BeST) is intended to provide a transparent and objective empirical framework for comparing digital self-triage solutions, including MDR-certified medical devices available in the EU. The present study evaluates the framework using the specific configurations and access pathways described below. Determining which system should be adopted also requires consideration of other factors, including feasibility and costs.

Case vignettes remain a practical evaluation approach, but their relevance depends on how well they represent real presentations. Seidel et al. developed the population-linked generation framework used to derive a human-manageable set of machine-generated candidate vignettes from large-scale routine triage data based on SmED (Strukturierte medizinische Ersteinschätzung in Deutschland)^11^. BeST did not use those machine-generated texts unchanged: before triage assessment, physicians thoroughly reviewed the raw texts, redacting or adding details where needed, splitting candidates into separate vignettes where appropriate, and removing near-duplicates to make the cases clearer and less ambiguous for assessment. This manuscript evaluates the methodology and presents the first BeST analysis, comparing two established digital triage technologies accessed outside their respective manufacturers’ MDR-regulated pathways — SmED via API and Infermedica Engine API via Microsoft Azure Healthcare Agent Service — with five OpenAI configurations prompted to conduct a structured triage assessment (gpt-4.1, gpt-5.4-2026-03-05, gpt-5.6-luna, gpt-5.6-sol, and gpt-6astra). The study did not evaluate either manufacturer’s certified medical device in its intended regulated configuration. The OpenAI configurations were not medical devices under the European Union (EU) Medical Device Regulation (MDR). For each case vignette, the reference was a preliminary consensus from five of the seven physicians in the study group, all employed at the Central Research Institute of Ambulatory Health Care in Germany (Zentralinstitut für die kassenärztliche Versorgung in der Bundesrepublik Deutschland; Zi). The non-LLM systems should not be treated as a single technical class. Infermedica has publicly described its general-public diagnostic system as a Bayesian-network implementation^12^. According to personal communication with its developers, this also applies to SmED, although other terms have been used in publications to describe how SmED arrives at its recommendations^7^. In a trial registration, the related SMASS self-triage technology was described as a computerised transparent neural network trained on evidence and specialist expertise, and its prospective real-world evaluation assessed safety in 2,543 emergency-care presentations^13,14^. Network-based approaches are well established in triage decision support and digital self-triage^15–17^. Accordingly, BeST compares two established non-LLM triage technologies evaluated through study-specific access pathways with five LLM configurations prompted to simulate structured triage assessments. The findings therefore characterize the evaluated implementations rather than the performance of the manufacturers’ certified medical devices.

## Results

### Reference distribution and coverage

Each vignette was assigned a winsorised population-representation weight derived from routine triage frequencies. The weighted and unweighted reference distributions are shown in Table 1. For each triage system, weighted coverage was defined as the share of the weighted vignette population for which the system produced an evaluable classification. It ranged from 98.1% to 100.0% of the known protocol population (Table 2); counts of evaluable and excluded vignettes are unweighted. Because the benchmark unit is a curated vignette rather than a recruited participant, demographic and clinical characteristics are represented through the direct population-linked weighting procedure rather than through a conventional participant baseline table. For readability, the SmED API and the Infermedica Engine API accessed via Microsoft Azure Healthcare Agent Service, both evaluated in study-specific configurations outside their respective MDR-regulated deployment pathways, are hereafter referred to as “SmED” and “Infermedica”, respectively. These designations refer exclusively to the evaluated configurations and not to the manufacturers’ certified medical devices.

**Table 1:** Distribution of Zi consensus labels.

| Zi consensus label | Vignettes, n | Unweighted percentage | Weighted percentage |
|---|---|---|---|
| Acute | 290 | 60.4 | 65.0 |
| Emergency | 95 | 19.8 | 16.2 |
| Not urgent | 95 | 19.8 | 18.8 |

**Table 2:** Frequency-weighted coverage by triage system; vignette counts are unweighted.

| Triage system | Evaluable, n | Excluded, n | Weighted coverage, % |
|---|---|---|---|
| SmED 5.4.11 (API) | 480 | 0 | 100.0 |
| Infermedica Eng.-API Azure | 469 | 11 | 98.1 |
| gpt-4.1 (structured) | 479 | 1 | 100.0 |
| gpt-5.4-2026-03-05 (structured) | 478 | 2 | 99.0 |
| gpt-5.6-luna (structured) | 480 | 0 | 100.0 |
| gpt-5.6-sol (structured) | 480 | 0 | 100.0 |
| gpt-6-astra (structured) | 480 | 0 | 100.0 |

### Overall and Class-specific performance

Figure 1 compares weighted macro-averaged sensitivity, specificity, and accuracy across systems, including 95% confidence intervals; full class-specific weighted estimates with confidence intervals are given in Table 3. Vignette counts remain unweighted. Corresponding unweighted overall and class-specific performance estimates are provided in Supplementary Tables S1 and S2. Weighted emergency sensitivity ranged from 51.3% (95% CI 35.5-66.3%) for gpt-4.1 to 99.9% (95% CI 99.7100.0%) for gpt-6-astra. The corresponding estimates for gpt-5.6-luna (96.7%), SmED (92.4%) and gpt-5.6-sol (90.4%) also had point estimates above 90%. Weighted acute sensitivity was highest for gpt-4.1 (82.0%), followed by gpt-5.4-2026-03-05 (70.6%); Infermedica had the lowest value (47.6%). Weighted not-urgent sensitivity was highest for Infermedica (64.3%), followed by gpt-5.6-sol (39.6%) and gpt-4.1 (38.1%). Weighted specificity ranged from 61.1% to 91.3% for emergencies, 45.9% to 79.0% for acute cases, and 74.9% to 100.0% for not-urgent cases; the full interval estimates are given in Table 3. Weighted exact three-class agreement (accuracy) was highest for gpt-4.1 (68.8%) and ranged from 54.4% to 64.8% for the other systems.

**Figure 1:**
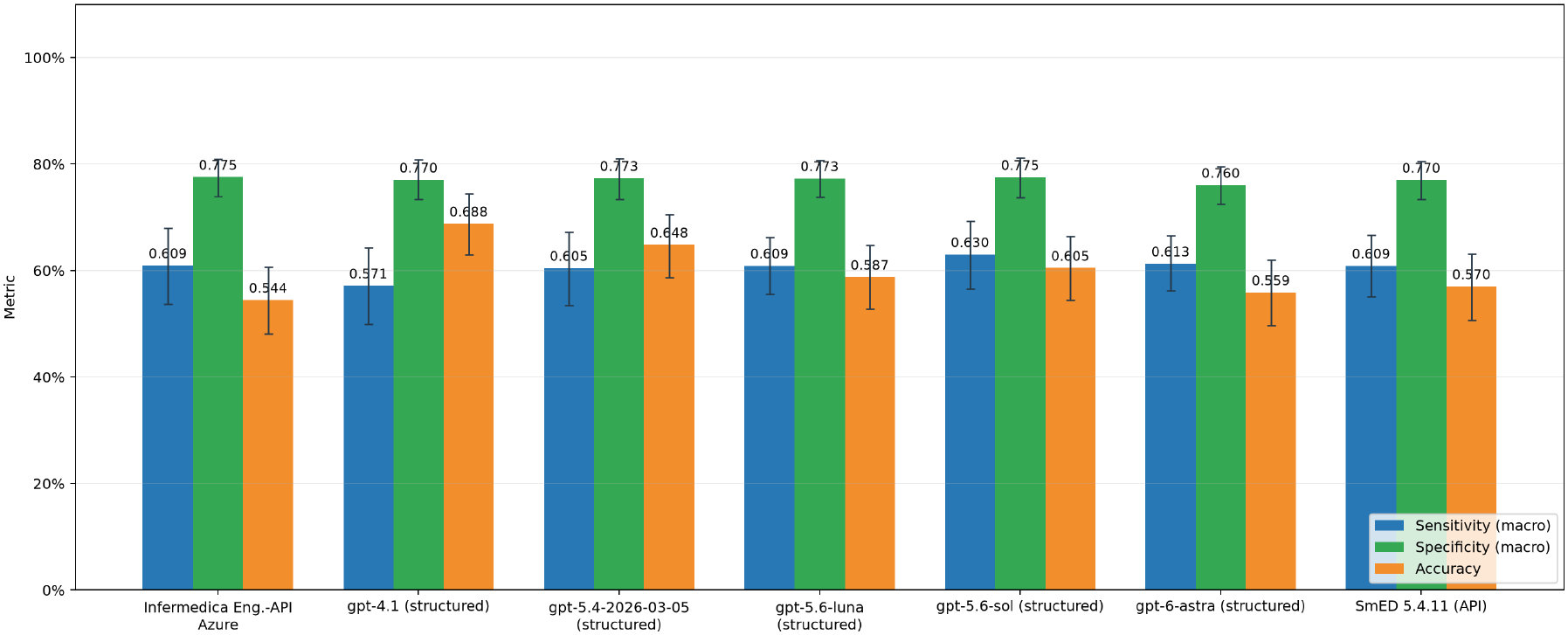
Winsorized frequency-weighted macro-averaged sensitivity, specificity, and accuracy by triage system.

**Table 3:** Winsorized frequency-weighted class-specific sensitivity and specificity against the Zi consensus; values are percentages with two-sided 95% bootstrap confidence intervals in parentheses. Evaluable vignette counts are reported in Table 2.

| Target category | Triage system | Weighted sensitivity, % (95% CI) | Weighted specificity, % (95% CI) |
|---|---|---|---|
| Emergency | Infermedica Eng.-API Azure | 70.9 (56.4-84.5) | 78.7 (73.1-84.2) |
|  | SmED 5.4.11 (API) | 92.4 (83.5-98.9) | 65.4 (58.8-71.9) |
|  | gpt-4.1 (structured) | 51.3 (35.5-66.3) | 91.3 (87.4-94.8) |
|  | gpt-5.4-2026-03-05 (structured) | 77.7 (63.3-90.8) | 78.6 (73.1-83.8) |
|  | gpt-5.6-luna (structured) | 96.7 (89.5-100.0) | 65.1 (58.6-71.4) |
|  | gpt-5.6-sol (structured) | 90.4 (79.3-99.9) | 68.6 (62.3-74.7) |
|  | gpt-6-astra (structured) | 99.9 (99.7-100.0) | 61.1 (54.3-67.8) |
| Acute | Infermedica Eng.-API Azure | 47.6 (39.9-55.5) | 79.0 (70.4-86.8) |
|  | SmED 5.4.11 (API) | 54.2 (46.3-61.8) | 69.8 (60.1-79.2) |
|  | gpt-4.1 (structured) | 82.0 (75.9-87.7) | 45.9 (35.8-56.5) |
|  | gpt-5.4-2026-03-05 (structured) | 70.6 (63.5-77.2) | 55.6 (45.0-65.8) |
|  | gpt-5.6-luna (structured) | 58.2 (50.7-65.7) | 67.6 (57.7-77.2) |
|  | gpt-5.6-sol (structured) | 59.0 (51.3-66.4) | 65.2 (55.1-74.8) |
|  | gpt-6-astra (structured) | 51.7 (43.9-59.6) | 66.9 (56.8-76.5) |
| Not urgent | Infermedica Eng.-API Azure | 64.3 (49.8-77.7) | 74.9 (68.6-80.9) |
|  | SmED 5.4.11 (API) | 36.0 (23.3-50.3) | 95.7 (92.7-98.3) |
|  | gpt-4.1 (structured) | 38.1 (24.8-52.2) | 93.8 (90.2-97.0) |
|  | gpt-5.4-2026-03-05 (structured) | 33.1 (19.6-47.3) | 97.7 (95.6-99.4) |
|  | gpt-5.6-luna (structured) | 27.7 (15.7-40.4) | 99.2 (97.9-99.9) |
|  | gpt-5.6-sol (structured) | 39.6 (25.6-53.8) | 98.7 (96.7-100.0) |
|  | gpt-6-astra (structured) | 32.1 (19.3-45.9) | 100.0 (100.0-100.0) |

Compared with SmED, gpt-6-astra had higher weighted emergency sensitivity (99.9% (95% CI 99.7100.0%) versus 92.4% (95% CI 83.5-98.9%)) and lower weighted emergency specificity (61.1% (95% CI 54.3-67.8%) versus 65.4% (95% CI 58.8-71.9%)). Weighted exact three-category agreement was 55.9% for gpt-6-astra and 57.0% for SmED. Thus, gpt-6-astra’s higher emergency sensitivity was accompanied by more frequent emergency classification of reference non-emergency cases.

### Multi-class classification

The winsorized frequency-weighted confusion matrices in Figure 2 make the pattern of overand under-triage compared to a preliminary expert consensus visible across all three categories. They should be interpreted with the system-specific weighted coverage in mind.

**Figure 2:**
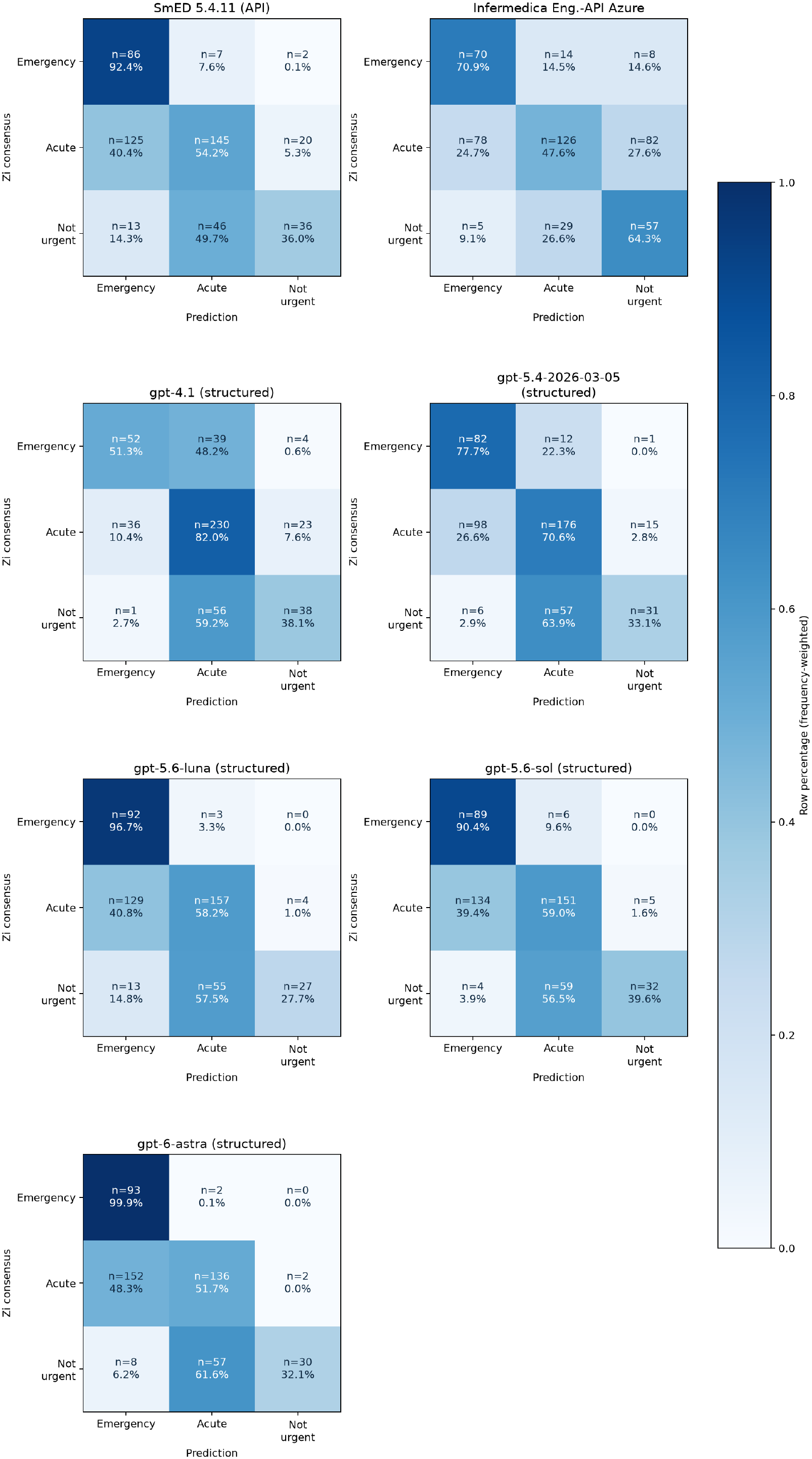
Winsorized frequency-weighted combined three-class confusion matrices for all currently included systems with raw counts.

### Actor-answer coverage sensitivity analysis

We computed actor-answer confidence scores for all 7 compared triage systems (SmED API 5.4.11, Infermedica Engine API Azure, gpt-4.1, gpt-5.4-2026-03-05, gpt-5.6-luna, gpt-5.6-sol, gpt-6-astra) under 7 system-specific patient-actor configurations. Each configuration covered 480 vignettes; together, the 7 collections comprised 3360 vignette records and 38,284 actor answers. At the main descriptive threshold of 0.90, 94.0% of the combined actor-answer mass, weighted by the final vignette weights, had a model-reported confidence score at or above the threshold. The sensitivity analysis across thresholds is shown in Table 4. These values describe model-based confidence scores that an answer is supported by the vignette.

**Table 4:** Vignette-frequency-weighted confidence-based actor-answer coverage sensitivity analysis by available system-protocol configuration (n = 480 vignette protocols per configuration); counts are unweighted.

| Triage system | Confidence threshold, % | Actor answers, n | Weighted coverage, % |
|---|---|---|---|
| SmED 5.4.11 (API) | 80 | 12105 | 97.6 |
| SmED 5.4.11 (API) | 90 | 12105 | 95.0 |
| SmED 5.4.11 (API) | 100 | 12105 | 36.3 |
| Infermedica Eng.-API Azure | 80 | 10180 | 94.3 |
| Infermedica Eng.-API Azure | 90 | 10180 | 91.2 |
| Infermedica Eng.-API Azure | 100 | 10180 | 23.0 |
| gpt-4.1 (structured) | 80 | 4706 | 96.8 |
| gpt-4.1 (structured) | 90 | 4706 | 94.7 |
| gpt-4.1 (structured) | 100 | 4706 | 27.8 |
| gpt-5.4-2026-03-05 (structured) | 80 | 3386 | 96.7 |
| gpt-5.4-2026-03-05 (structured) | 90 | 3386 | 94.9 |
| gpt-5.4-2026-03-05 (structured) | 100 | 3386 | 29.1 |
| gpt-5.6-luna (structured) | 80 | 3891 | 97.0 |
| gpt-5.6-luna (structured) | 90 | 3891 | 94.5 |
| gpt-5.6-luna (structured) | 100 | 3891 | 23.1 |
| gpt-5.6-sol (structured) | 80 | 2301 | 97.2 |
| gpt-5.6-sol (structured) | 90 | 2301 | 94.3 |
| gpt-5.6-sol (structured) | 100 | 2301 | 24.0 |
| gpt-6-astra (structured) | 80 | 1715 | 98.1 |
| gpt-6-astra (structured) | 90 | 1715 | 96.7 |
| gpt-6-astra (structured) | 100 | 1715 | 14.7 |

For the SmED interactions, the weighted mean actor-answer confidence score was 0.966, compared with 0.936 for Infermedica. At the primary confidence threshold of 0.90, weighted actor-answer coverage was 95.0% for SmED and 91.2% for Infermedica; coverage was also higher for SmED at each of the other reported thresholds (Table 4).

Across the OpenAI configurations, the number of recorded actor answers was 4,706 for gpt-4.1, 3,386 for gpt-5.4-2026-03-05, 3,891 for gpt-5.6-luna, 2,301 for gpt-5.6-sol, and 1,715 for gpt-6-astra (Table 4). The number of recorded answers was 63.6% lower for gpt-6-astra than for gpt-4.1. The decline in answer counts was not uniform across configurations: gpt-5.6-luna had more recorded answers than gpt-5.4-2026-03-05.

### Exploratory manual protocol review

The exploratory manual review covered all 31 trajectories in which SmED or Infermedica in their study-specific non-MDR-regulated configurations assigned a lower urgency than the reference emergency category: 22 Infermedica and nine SmED trajectories from 28 distinct vignettes. Three reviewer entries per trajectory yielded 93 ratings. Complete entries unanimously identified a documented deviation in 14 trajectories (5x SmED, 9x Infermedica) and no deviation in 12 trajectories (4x SmED, 8x Infermedica); 3 (all Infermedica) were unanimously rated not assessable. Two (both Infermedica) had discordant ratings. Reviewer comments described omitted vignette information, difficulties mapping vignette information to response options (especially when it was only implied), and inaccurate symptom timing. These observations did not establish whether a deviation caused the final undertriage classification. The review did not change the primary classifications or performance estimates (Supplementary Tables S3 and S4).

## Discussion

This is a feasibility study describing a potential evaluation process of complex self-triage systems in which we use a representative and physician-curated set of patient vignettes to create recommendations from self-triage systems which are then benchmarked against a preliminary expert consensus. The information contained in the patient vignettes was used to generate automated queries to the included self-triage systems. The aim of this paper is to demonstrate how self-triage systems could be benchmarked against a future medical gold standard, and what information could be generated by this benchmarking process. The primary contribution is methodological: the results obtained against the preliminary consensus illustrate the types of outcomes this framework can produce. They represent an initial benchmark iteration, not a completed assessment of the included systems.

The triage systems differed in how they balanced recognition of emergency presentations against overclassification, and performance patterns varied across urgency categories. The OpenAI configurations prompted to conduct a structured triage assessment and SmED generally had low weighted sensitivity for not-urgent cases (27.7%-39.6%) while retaining high weighted specificity for that category (93.8%100.0%), a pattern consistent with sparing use of the not-urgent label and a tendency toward higherurgency recommendations. Infermedica differed: it had the highest weighted not-urgent sensitivity (64.3%) but lower weighted not-urgent specificity (74.9%). The current comparison against the preliminary expert consensus is an evaluation of agreement with that reference, not proof of clinical effectiveness or safety, so a further revision of this reference with medical experts is needed, to become a standard for safety evaluation.

This heterogeneity in performance, rather than a single acuity gradient, is in line with much of the cited symptom-checker literature. Semigran et al. reported correct triage advice in 80% of emergency cases, 55% of non-emergency cases, and 33% of self-care cases, while Hill et al. likewise found higher accuracy for emergency (63%) and urgent care (56%) than for non-urgent care (30%) or self-care (40%)^1,18^. A five-year follow-up found no overall improvement and reported that the evaluated apps missed more than 40% of emergencies, although performance varied widely between products^19^. More recently, a systematic review found moderate but highly variable overall accuracy for symptomassessment applications and a narrower moderate range for LLMs; across technologies, emergency cases were generally recognised better than self-care cases, and LLMs rarely recommended self-care^20^. BeST reproduces the difficulty of recognising cases that can remain outside urgent care for SmED and the five OpenAI configurations, whose weighted not-urgent sensitivities ranged from 27.7% to 39.6%; Infermedica was an exception (64.3%). The OpenAI configurations also varied markedly in weighted emergency sensitivity, from 51.3% for gpt-4.1 to 99.9% for gpt-6-astra, reinforcing that a result from one structured GPT configuration is not representative of the others. Direct numerical comparisons remain limited by different vignette sets, category mappings, interaction protocols, and outcome definitions. For example, Ceney et al. assessed the safety of disposition advice rather than exact class agreement and found safety to be lower for emergency than for non-urgent cases^2^.

The randomised study by Bean et al. adds an important user-level qualification to this literature. Although the LLMs performed well when assessed without participants, people receiving LLM assistance identified relevant conditions in fewer than 34.5% of cases and chose the correct disposition in fewer than 44.2%, with no advantage over the control condition^21^. This gap between standalone and assisted performance is consistent with the interaction-sensitive design of BeST: the present benchmark evaluates a specified patient-system exchange rather than treating the LLM as a questionanswering engine operating on a complete vignette. However, BeST uses a LLM-based simulated patient-role actor and physician consensus rather than members of the public and prospective clinical outcomes; the results therefore cannot determine whether the interaction protocol improves or impairs real-world decisions.

When patient safety is valued above efficiency, failure to recognise a reference emergency is a key benchmark concern. Weighted emergency sensitivity was highest for gpt-6-astra (99.9%), followed by gpt-5.6-luna (96.7%), SmED (92.4%) and gpt-5.6-sol (90.4%); gpt-4.1 had the lowest value (51.3%). This ordering is an agreement-based safety signal only: relative to the preliminary Zi consensus, the former configurations produced fewer weighted false-negative emergency classifications in these vignettes. It does not establish clinical safety, because the reference is preliminary, the outcomes are vignette-based, and the analysis does not include prospective patient outcomes. Nevertheless, a lower weighted emergency sensitivity warrants scrutiny before any safety-critical deployment of a product.

The weighted emergency results show a safety-efficiency trade-off rather than a single ranking. The configurations with the highest weighted emergency sensitivity had weighted emergency specificity of 61.1%-68.6%, meaning that more non-emergency vignette mass was assigned to the emergency category. gpt-4.1 showed the converse profile, with the highest weighted emergency specificity (91.3%) and the lowest weighted emergency sensitivity (51.3%). Infermedica combined intermediate weighted emergency sensitivity (70.9%) and specificity (78.7%). Such descriptive trade-offs identify priorities for prospective validation, future development and even the very role of self-triage. Since safety should not be traded for efficiency, the sensitivity-specificity relationship argues for sequential testing using different methods (including i.e. human medical professionals), to reduce avoidable utilization of emergency care without compromising patient safety.

The comparison between the two established non-LLM triage technologies, evaluated through studyspecific access pathways, and five OpenAI configurations does not reveal a simple separation between these technological groups. gpt-4.1 had the highest weighted acute sensitivity and weighted exact three-class agreement, gpt-6-astra had the highest weighted emergency sensitivity, and Infermedica had the highest weighted not-urgent sensitivity. No system therefore dominated all categories. Differences among the OpenAI configurations were also substantial, particularly for weighted emergency sensitivity (51.3%-99.9%), showing that a result from one configuration cannot be treated as representative of all LLMs or of one model family. These comparisons do not show regulatory or clinical equivalence: the benchmark does not assess intended-use conformity, quality management, risk controls, software-change control, post-market surveillance, or real-world patient outcomes. Published LLM findings likewise vary by model and protocol, ranging from a tendency towards overtriage in emergency-medicine vignettes to substantial under-triage in a structured consumer-health evaluation^3,4^. The present results belong to the exact model snapshots, prompts, and interaction workflows tested.

Across the evaluated OpenAI configurations, the results suggest a shift towards the precautionary triage profile. Weighted emergency sensitivity increased from 51.3% for gpt-4.1 to 77.7% for gpt5.4-2026-03-05 and exceeded 90% for both gpt-5.6 configurations and gpt-6-astra, while emergency specificity was lower in these configurations. With emergency sensitivity of 99.9% and specificity of 61.1%, gpt-6-astra extended this pattern towards more frequent emergency recommendations. This is consistent with greater prioritisation of avoiding missed reference emergencies, accompanied by increased over-triage relative to the preliminary consensus. The higher emergency sensitivity did not translate into higher overall agreement: exact three-category agreement was 55.9% for gpt-6-astra, compared with 68.8% for gpt-4.1.

This pattern is compatible with an evolution towards more precautionary triage behaviour within the evaluated model family, but the cross-sectional comparison does not establish a longitudinal or uniformly monotonic improvement. OpenAI likewise reports a higher length-adjusted HealthBench Professional score for gpt-6-astra than for gpt-5.6-sol (63.4% versus 60.5%), describing this as an improvement on clinician-facing health tasks^22,23^. This external finding is directionally compatible with a generation effect, but HealthBench Professional deliberately enriches difficult cases and uses a different task, harness, sampling scheme, and rubric-based outcome; its scores are therefore not directly comparable with BeST results. The BeST findings should accordingly be interpreted as generation-specific changes in the tested triage profile, not as evidence of a general or longitudinal improvement in clinical safety. It also cannot distinguish improved recognition of clinical urgency from a greater propensity to recommend emergency care. Differences in training, post-training, inference behaviour, and the simulated patient-system interaction may all contribute. Similarity to SmED therefore concerns the observed benchmark error profile; it does not establish equivalent clinical safety, regulatory status, or real-world effectiveness.

Our study also sheds light on another aspect of the compared triage systems that is likely to be relevant for implementation as self-triage increasingly becomes part of digital access points to national health care systems. In Germany for example government is preparing a law^9^ which would create a standardised digital access point to medical care focusing on providing a central tool for self-triage which may be accessed by phone, web or app. This approach has been tested in England where the NHS App in England also supports access to the personal general practitioner (GP), access to clinical data and prescriptions, as well to a central digital self-triage in case of a suspected emergency^24^.

For England it has been shown that a two-stage approach in triaging patients’ demands might be useful with a sensitive self-triage followed by a more specific triage by medical professionals^25^. In our study the number of questions asked by the triage system varies considerably between the two structured systems and the LLM (Table 4). Also, more recent OpenAI GPT models required less questions than earlier ones. Placing a short but sensitive triage up front might therefore be a strategy to select patients which need to receive a second-level triage by health professionals. Complex questioning of self-triage systems has been criticised by GPs in Germany when introducing digital concierge systems in their surgeries. Because the introduction of digital communication portals in general practice is still a new concept there is little published evidence on how the complexity of structured self-assessments affects access to care. Ease of access, however, is a theme which is why fast decision making tools are likely to be favored over complex questionaries^26^.

The current implementation and analysis has important limitations. The reference is preliminary, results are based on a finite vignette set, and weighted estimates depend on the similarity-based reassignment and the pragmatic Winsorisation rule. The machine-generated candidate set was designed to retain an explicit link to the underlying triage population^11^, and the final BeST vignettes were substantially modified through physician enrichment, splitting, and deduplication. The initial statistical derivation of candidate vignettes from SmED routine protocols embeds a SmED-related source bias in the benchmark: case selection and available clinical information are shaped by what this system elicits and records. Physician curation can mitigate this source dependence but cannot fully eliminate it or ensure coverage of information relevant to other triage approaches. A further limitation concerns the configurations of the established triage technologies evaluated in this study. SmED was accessed through an API outside its standard MDR-regulated deployment pathway, while the Infermedica Engine API was accessed via Microsoft Azure Healthcare Agent Service using a study-specific implementation. Neither system was evaluated through the manufacturer’s standard regulated deployment pathway. In particular, the evaluated Infermedica configuration was not the manufacturer’s certified Medical Guidance Platform (MGP). The findings therefore apply to the specific implementations and interaction workflows used in BeST and should not be interpreted as a direct evaluation of the performance or clinical safety of the manufacturers’ certified medical devices in their intended deployment settings.

To preserve the link between candidate vignettes and triage population, population weights were re-evaluated after each textual redaction and reassigned where necessary as part of the iterative curation process rather than transferred unchanged from the candidate set. The resulting estimates should not be read as design-based population estimates: the protocol corpus is incomplete, the match threshold and percentile bounds are not empirical prevalence models, and semantic match quality still requires independent validation. The project retains the unweighted vignette counts and the uncapped representation weights, so that future protocol additions and alternative weighting rules can be assessed transparently.

The information sequence is a further potentially material source of bias. For a fully specified Bayesian network, the posterior conditional on an identical complete set of observations is invariant to the order in which those observations are entered. In an implemented self-triage dialogue, however, the question path is adaptive: early answers determine which subsequent questions are asked, whether the interaction ends early, and which information is never elicited. In BeST, the LLM-based patient-role actor additionally generates each answer in the context of the preceding dialogue. The order in which vignette information becomes available can therefore influence the questions presented and the final recommendation, even when the underlying vignette is unchanged. We did not randomise or formally quantify these order effects, so the estimates apply to the exact interaction sequence used. Future work should further develop the patient-role actor and evaluate it with repeated, counterbalanced information orders and predefined response variants; this is necessary to reduce sequence-dependent bias and to distinguish product performance from artefacts of the simulated patient interaction.

The exploratory manual review identified documented deviations in 14 of 31 reference-emergency undertriage trajectories. Reviewer comments described relevant information that was not communicated despite opportunities to add it, difficulties mapping symptoms to available response options, and inaccurate representation of symptom timing. These findings indicate that benchmark performance reflects the combined vignette, patient actor, interface, and triage-system workflow. They are compatible with interaction-related information loss but do not establish a causal effect of question order. Because the review was restricted to undertriage cases from two systems and was not blinded, it cannot estimate overall actor error rates, establish comparative actor reliability across systems, or validate the five OpenAI triage configurations. The twelve trajectories without a documented deviation do not establish complete information transfer or clinical appropriateness of the final recommendation. Nor do the identified deviations show how much undertriage was attributable to the simulated interaction: their effects on final recommendations were not systematically tested.

Some reviewer concerns reflected the actor prompts’ explicit assumption that specified categories of unmentioned information could be treated as absent. A response may therefore follow the prompt while inadequately representing clinical uncertainty. Conversely, a clinically plausible inference, such as an associated symptom not stated in the vignette, is not by itself evidence of an actor error. Future evaluations should distinguish departures from vignette content or operational rules from limitations of those rules, interface constraints, and vignette ambiguity. Prespecified corrected replays and repeated runs should assess whether these issues change final recommendations, without retrospectively replacing the primary benchmark results.

The confidence-based actor-answer coverage analysis provides an additional diagnostic of the simulated interaction, but it is not an independent validation of actor fidelity: confidence scores are produced by the same model that selected the answer. The preceding vignette-generation study reported manual review without relevant abnormalities^11^, but that finding does not establish fidelity in the present system-specific dialogues. The confidence-based estimates and the targeted manual review are complementary descriptive diagnostics; neither replaces a representative assessment of actor fidelity or a repeated-run stability analysis.

The higher actor-answer confidence and confidence-based coverage in SmED interactions than in Infermedica interactions may partly reflect the provenance of the vignettes. The source data consisted of routine SmED triage protocols, so the information represented in the vignettes is likely to align more closely with the questions and answer options elicited by SmED. Despite substantial physician curation, this alignment may make it easier for the patient-role actor to identify explicit vignette support for answers to SmED questions, whereas Infermedica may elicit information that was less consistently captured in the source protocols. This provides a plausible explanation for the observed difference, but the contribution of source alignment was not separately tested. Differences in question paths, answer options, and system-specific actor prompts may also contribute. Higher confidence in this setting therefore does not by itself demonstrate more accurate actor answers or superior clinical performance; it may also indicate an advantage arising from the benchmark’s source material. This limitation extends beyond confidence-based actor-answer coverage to the interpretation of comparative benchmark outcomes. Unequal availability of information relevant to different systems may influence their final classifications and agreement with the reference. The direction and magnitude of any resulting effect on comparative performance have not been separately quantified.

The comparison across OpenAI generations also suggests that shorter simulated dialogues can coexist with higher emergency sensitivity. In particular, gpt-6-astra required substantially fewer recorded actor answers than gpt-4.1 while identifying a larger proportion of reference emergencies. This pattern may reflect more selective questioning or earlier decisions to recommend emergency care. The latter interpretation is consistent with the lower emergency specificity of gpt-6-astra, so fewer questions should not automatically be interpreted as more efficient clinical information gathering. The pattern was not uniform across configurations, as illustrated by the higher answer count for gpt5.6-luna than for gpt-5.4-2026-03-05. Recorded answer counts do not capture question complexity, response burden, or interaction time, and higher emergency sensitivity is an agreement-based safety signal rather than evidence of improved clinical safety. Future evaluations should assess whether shorter dialogues reduce user burden while retaining adequate information for safe and appropriately specific triage recommendations.

The preliminary Zi consensus is a reference standard for this analysis, not a definitive ‘gold standard’. Because the representative-vignette redaction and consensus process were carried out by seven physicians with general knowledge of SmED, the process could have favoured SmED despite blinding to its vignette-level results. This possible source of bias should be considered when interpreting comparative agreement with the Zi label. This potential reviewer-related bias is distinct from the source dependence introduced during vignette generation. The planned involvement of medical specialty societies is intended to broaden the review of both vignette content and reference classifications, including scrutiny of assumptions inherited from the source system.

The use of a standardised system prompt for LLMs does not represent the wide range of ordinary real-world interactions with these models. We nevertheless consider it necessary in this benchmark because it establishes a controlled and reproducible interface that can be compared fairly with structured medical questionnaires. This design therefore evaluates a specified LLM triage configuration rather than the unconstrained use of a general-purpose LLM.

As the next step in BeST, Zi aims to open the consensus process to medical specialty societies, with the goal of developing a more broadly supported expert reference standard for vignette evaluation. We will also extend the patient-role actor beyond the current vignette-faithful response model to represent a broader range of patient characteristics and communication needs, including limited German-language proficiency and reduced cognitive capacity, among others. These developments are intended to make future benchmark scenarios more representative of the populations for whom self-triage tools are used. BeST is intended as an ongoing programme of vignette refinement, broader clinical consensus formation, and repeated evaluation. The present vignette set, reference classifications, and comparative results therefore represent a preliminary stage in this process. Future benchmark iterations will document revisions explicitly so that changes in results can be interpreted in relation to changes in the vignettes, reference standard, system configurations, and evaluation workflow.

## Methods

### Study design and vignette set

The complete study workflow is summarised in Figure 3. The population-linked generation phase follows Seidel et al.^11^, whereas physician curation, consensus formation, system execution, and comparative analysis constitute the BeST workflow reported here.

**Figure 3:**
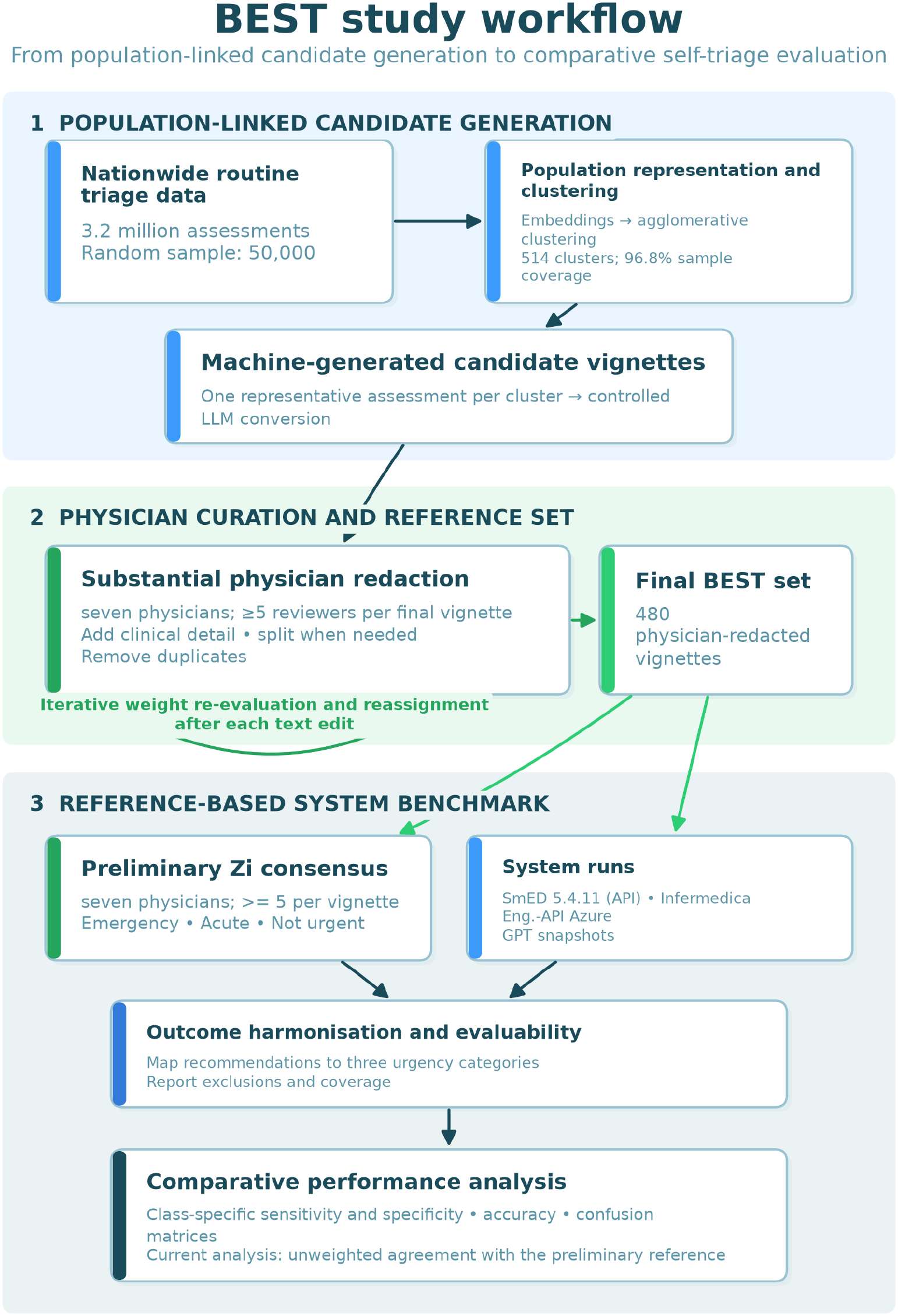
Study workflow from routine triage data and machine-generated candidate vignettes through iterative physician curation and weight reassessment, preliminary consensus formation, system execution, and comparative performance analysis.

This comparative vignette-based accuracy study evaluated triage-system classifications against a preliminary physician consensus. The population-linked candidate-vignette generation followed the framework developed by Seidel et al.^11^ and used routine triage data collected by SmED (via telephone, self-triage or at joint contact points of the outpatient emergency care service and hospital emergency departments) between 1 November 2023 and 31 October 2024. An initial sample of 50,000 assessments was drawn randomly from the eligible source population; no participants were recruited for this study. This sample size was used because clustering algorithms become increasingly computationally demanding as the number of data points grows. The given size was a pragmatic compromise between population coverage and computational feasibility.

The representative assessments were enriched using a two-phase simulated-annealing procedure. This automated generation and enrichment phase produced 514 population-linked candidate vignettes. Population weighting was referenced to the full 2025 routine-triage protocol population without applying the initial sampling fraction.

Before the final urgency rating and consensus process, achieved by a seven-member pool of physicians from different medical specialties, six of them substantially redacted the candidate set. The physicians did not know the vignette-level SmED classifications while forming the consensus, but had general knowledge of SmED as a triage system. Every final vignette was reviewed and redacted by at least two physicians. The physicians added clinical details when needed or rephrased to improve clarity, split a candidate into separate vignettes when distinct presentations required separate assessment, and removed practically identical or otherwise unsuitable candidates. This physician-led medical editorial transformation produced the final 480 BeST vignettes and was intended to create clearer, sufficiently specified, and non-duplicative cases for rating while removing residual contextual links to the source records. The resulting final vignettes contained no direct identifiers or linkable case identifiers and were non-identifiable in the study data. These transformed final vignettes, rather than the original machine-generated candidates, constitute the benchmark set analysed in this manuscript. Physician curation and consensus formation took place from Dezember 2025 through July 2026, followed by system evaluation in August 2026.

Weight maintenance was integrated into this curation cycle. After each textual redaction or structural change to a vignette, its population weight was re-evaluated and reassigned where necessary before the vignette proceeded to final rating. Redaction and weight reassessment therefore formed an iterative process rather than two independent sequential steps.

For the final reassignment, each assessment *i* from the 2025 routine-triage population was compared with each final vignette *j* using cosine similarity *s_ij_* between their Sentence-Transformer text embeddings. At the selected similarity cutoff *c* = 0.85, the match set for assessment *i* was *M_i_*(*c*) = {*j* ∶ *s_ij_* ≥ *c*} and its size was *m_i_*(*c*) = |*M_i_*(*c*)|. For assessments with at least one match, a fractional weight was allocated equally across all matching vignettes:

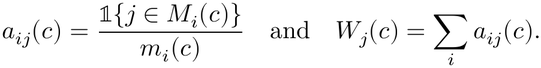

Thus, every covered assessment contributes a total weight of one across its matching vignettes, while a vignette without a match receives weight zero. At *c* = 0.85, the raw allocated weights covered 90.7% of population assessments; 34 of the final vignettes had raw weight zero. A one-way sensitivity analysis evaluated cutoffs from 0.80 to 0.99 in increments of 0.01, reporting both the proportion of distinct assessments with at least one match and the total match mass. The cutoff was selected as the practical trade-off between maintaining coverage and limiting multiple assignments; the sensitivity analysis is descriptive and does not validate semantic equivalence of matched texts.

The protocol corpus does not encompass all real-world presentations, and the benchmark also contains medically curated vignettes that may have no match in the currently available questionnaires. We therefore used two-sided Winsorisation of the raw representation weights before performance analysis. Let *L* = *Q*_0.20_(*W* ) and *U* = *Q*_0.80_(*W* ) be the 20th and 80th percentiles across all final vignettes, including zero weights. The Winsorised weight was

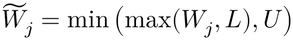

It was subsequently normalised to the known protocol population:

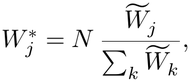

where *N* is the number of assessments in that population. This assigns every vignette at least *L*, limits the influence of individual high-weight vignettes, and preserves the ordering and weight differences within the central 60% of the empirical distribution. Weight trimming or Winsorisation commonly trades a reduction in weight variability against introduced bias; the choice of bounds is consequential and should not substitute for improving the underlying weighting process^27^. For this benchmark, that limitation was preferable to equal vignette weights, which would discard all observed differences in protocol representation, while preventing any medically curated vignette from being completely excluded. The percentile bounds remain a pragmatic regularisation choice, the additional lower-tail weight is not an empirical prevalence estimate, and neither the transformation nor the subsequent normalisation can recover presentations missing from the protocol corpus. Accordingly, weighted results are interpreted as a sensitivity analysis based on regularised representation weights, not as design-based population estimates.

No formal subgroup fairness analysis was planned for this proof-of-concept benchmark. The vignetteconstruction framework achieved approximately 98% coverage of the underlying random source sample in the preceding evaluation^11^, and the final weights were linked directly to the full source population. This design reduces concern about omission caused by the initial sample, but does not establish that all residual demographic or clinical subgroup biases are absent; such analyses are planned for future benchmark updates.

#### Vignette flow

We generated 514 machine-generated candidate vignettes, added 7 through splitting, and excluded 37 for several reasons (e.g., nearly identical vignettes), yielding 484 vignettes. Each vignette went through an initial annotation process and was then thoroughly reviewed by at least 2 physicians; reviewers had access to the vignettes and pre-defined answer options and wereblinded to system outputs during content revision and consensus rating. During the curation process we added 10 more case vignettes as variations, and we excluded 14 vignettes primarily focusing on psychiatric symptoms because we intend to edit them together with experts. The final dataset for this analysis comprised 480 vignettes.

#### Reference standard

The reference standard was established by seven licensed physicians, six of whom were board-certified in different clinical specialties, including general practice, paediatrics, internal medicine and anaesthesia, and had at least five years of clinical experience. Three of them held the additional qualification in emergency medicine. One physician had not yet completed specialist training but a strong background in the provision of emergency care. From this pool, five physicians independently rated each vignette via a software tool. Disagreements were resolved in consensus conferences, each involving at least five of the seven study group physicians; therefore, the group composition varied across vignettes. This preliminary physician consensus was selected as the initial reference because sufficient medical expertise was available for an initial assessment, whereas consultation with medical specialty societies could not be completed within the project timeline. Broader specialty-group consultation and a revised consensus process are planned for a subsequent version of BeST.

### Ethics and consent

Formal ethics committee approval was not required for this vignette-based study.

The data used for the data-driven vignettes^11^ consist of pseudonymized individual-level social data that were lawfully collected and stored in accordance with Section 285(3) in conjunction with Section 285(1) No. 6 of the German Social Code, Book V (SGB V), and may be processed for quality assurance purposes. The data are owned by the Associations of Statutory Health Insurance Physicians (ASHIPs). On the basis of contractual data processing agreements pursuant to Section 80 of German Social Code, Book X (SGB X) in conjunction with Article 28 of the General Data Protection Regulation (GDPR), the Central Research Institute of Ambulatory Health Care in Germany (Zi) processes these data in pseudonymized form for quality assurance. Data transfer and processing were reviewed for compliance with applicable data protection regulations. Access to the data was restricted to authorized personnel.

The system-evaluation inputs were the final physician-redacted and medically reviewed vignettes described above, not raw routine records or identifiable patient data. No direct identifiers or linkable case identifiers were transmitted to the evaluated systems.

The physician evaluation of the vignettes was conducted offline and did not involve patient interventions. It used non-identifiable vignettes derived from secondary data originally assembled for the work of Seidel et al.^11^, rather than real patient encounters or live patient data. The study did not involve patient interventions or assess adverse events in clinical use.

### Triage systems and LLM evaluation

The evaluated systems were Structured Medical Initial Assessment in Germany (Strukturierte medizinische Ersteinschätzung in Deutschland; SmED), the Infermedica Engine API accessed via Microsoft Azure Healthcare Agent Service, and five structured OpenAI configurations: gpt-4.1, gpt5.4-2026-03-05, gpt-5.6-luna, gpt-5.6-sol, and gpt-6-astra. SmED and Infermedica were selected as established digital triage technologies, while OpenAI was selected as a leading LLM provider. The initial proof-of-concept paper was not intended to be exhaustive; additional systems, including openweight models, are planned for future analytical updates. SmED 5.4.11 was accessed through a non-certified API pathway, while Infermedica Engine API was accessed via Microsoft Azure Healthcare Agent Service using a study-specific implementation. The implementation was developed by the study team and was not reviewed or validated by Infermedica. Neither evaluated configuration represented the manufacturer’s certified medical device in its intended regulated deployment setting. The Infermedica Engine API was accessed through a paid subscription to Microsoft Azure Healthcare Agent Service using a study-specific implementation. This implementation was not Infermedica’s certified Medical Guidance Platform (MGP), a Class IIb medical device under the European Union

Medical Device Regulation (MDR). Neither system was evaluated through its manufacturer’s standard MDR-regulated deployment pathway. The OpenAI configurations were not medical devices. No specific manufacturer approval or endorsement of this study was obtained from Infermedica. The OpenAI configurations were accessed through the OpenAI API on a pay-per-use basis. These access arrangements describe the technical and licensing basis of the evaluation and do not imply sponsorship, participation in study design, or endorsement of the results by any provider. Infermedica’s publicly described general-public diagnostic system uses a Bayesian-network methodology^12^. SmED has been described as a red-flag-based, neural-network-structured medical rule set^7^; the associated SMASS technology is described as a computerised transparent neural network^13,14^. Meer’s unpublished presentation further characterises SmED’s use of prior-triage information and posterior probability updating as a “Bayes network”^28^. This is a first-party explanation of the functional logic, not a peer-reviewed technical specification; it therefore supports, but does not definitively establish, a Bayesian-network classification.The large language model (LLM) benchmark used separate triage-system and patient roles, fresh sessions for each vignette, and structured JavaScript Object Notation (JSON) exchanges. The Supplementary Information presents the LLM workflow and English translations of its German operational prompts as an example illustrating the patient role-play approach. The remaining system-specific workflows for SmED and Infermedica are internal, unpublished implementations and are not included in the publicly available materials.

The system prompt was manually developed to make the LLM and triage-system results as comparable and reproducible as possible and to support standardised comparisons across LLM generations. It was fixed before the evaluation. The evaluation vignettes were kept separate from prompt development and were not used for post hoc prompt optimisation.

The analysis was conducted in August and September 2026. The benchmarked SmED version via API was 5.4.11, and the evaluated Infermedica configuration was designated as Infermedica Engine API Azure, accessed via Microsoft Azure Healthcare Agent Service. The evaluated OpenAI configurations were gpt-4.1, gpt-5.4-2026-03-05, gpt-5.6-luna, gpt-5.6-sol, and gpt-6-astra. Wherever technically possible, all configurations used the same prompts and workflow.

#### Interaction-sequence reproducibility

The primary analysis used one run per model-vignette combination in fresh, separate patient and triage sessions, without aggregation across repeated outputs. The patient actor was always simulated by gpt-5.4-2026-03-05 and instructed to answer each question in JSON using the vignette and the applicable system-specific response rules. The SmED and Infermedica actor prompts explicitly treated the vignette as complete for specified categories, including symptoms, comorbidities, risk factors, recent procedures, and pregnancy, and required a negative answer when such information was absent and an explicit negative option was available. Other unresolved responses were handled according to the available options: SmED generally used an unanswered response, whereas Infermedica used Unclear where available, an empty selection, or the closest available single-choice option. These instructions describe intended behaviour, not verified fidelity of every answer. Runs terminated on triage completion or predefined exchange limits/failure criteria. The OpenAI configurations were run with the standard OpenAI API settings; no additional temperature, seed, sampling, or other inference-parameter overrides were applied. The aggregate results data and analysis code will be published on GitHub after publication. English translations of the operational prompts for the LLM workflow are provided in the Supplementary Information as an example illustrating the patient role-play approach. The scripts used to conduct the role-play interactions are internal implementations and will not be made publicly available. Because order robustness and repeated-run stability were not systematically assessed, results apply to the exact information sequence and single-run configuration used.

#### Actor-fidelity diagnostics

For each question asked by the triage system, a separate LLM call assessed whether the patient actor’s corresponding answer was supported by, contradicted by, or compatible with the information in the case vignette. The evaluator received the current questionanswer pair and, where available, the immediately preceding pair from the interaction history. Using the same OpenAI model as the patient-role actor (gpt-5.4-2026-03-05), it assigned a confidence score ranging from 0 to 1 and generated an evidence field containing either a brief supporting excerpt from the vignette or a concise justification. These assessments were conducted separately from the actor-triage-system interaction and were analysed as confidence-based coverage diagnostics rather than independent validation of answer correctness.

#### Exploratory manual protocol review

We reviewed all trajectories in the current analysis dataset for which the Zi reference was Emergency and either SmED 5.4.11 via API or Infermedica Engine API Azure returned Acute or Not urgent. Missing system classifications were not counted as undertriage. This yielded 31 trajectories from 28 distinct vignettes. Reviewers had access to the vignette, recorded questions, response options, actor answers, and system results; the review was not blinded. The worksheet requested separate assessments and contains three reviewer entries per trajectory (R1-R3). Some entries explicitly document subsequent joint discussion, so the retained ratings are not assumed to be fully independent pre-discussion assessments. Reviewers recorded a deviation, no deviation, or inability to assess. An inability-to-assess rating required an explanation. At trajectory level, incomplete entries took precedence over disagreement, and a substantive outcome was assigned only when all three complete entries agreed. No majority-vote adjudication was applied. Counts were unweighted, and repeated ratings of the same trajectory were not treated as independent observations. Free-text explanations and supplementary worksheet remarks were examined descriptively, without a formal thematic coding or attribution of causal effects. The review did not alter the primary system classifications, reference labels, or performance estimates. An isolated corrected assessment mentioned in the review notes was not part of the primary analysis or a systematic replay experiment. Further procedural details and examples are provided in the Supplementary Information.

### Outcome harmonisation

All recommendations were recorded into three mutually exclusive clinical-urgency categories: **Emergency**, **Acute**, and **Not urgent**. The rule-based mapping is specified in Table 5. SmED and the LLM configuration both supplied a time-to-treatment (TTT) and point-of-care (POC) recommendation, which were combined using the same rule. Infermedica Engine API Azure supplied a single triage-level value, which was mapped directly. Its developer documentation distinguishes consultation_24, recommending that the patient sees a doctor within 24 hours, from consultation, for which an appointment may be scheduled if medical evaluation is required^29^. BeST was designed to assess clinical urgency rather than the recommended level of care. Accordingly, consultation_24 was mapped to Acute, whereas consultation and self_care were both mapped to Not urgent. The distinction between self-care and non-urgent physician consultation was outside the scope of this three-category urgency assessment. For the LLM configurations, *telemedical consultation* was mapped to telemedicine. Telemedicine recommendations were classified according to TTT: emergency as Emergency; immediate or within 24 hours as Acute; and not urgent as Not urgent.

**Table 5:** Rule-based mapping of system recommendations to the three harmonised clinical-urgency categories. Infermedica defines consultation_24 as seeing a doctor within 24 hours and consultation as arranging an appointment when medical evaluation may be required^29^. For the LLM, *telemedical consultation* was mapped to telemedicine.

| Harmonised category | SmED and LLM: TTT and POC rule | Infermedica triage level |
|---|---|---|
| Emergency | TTT: emergency or immediate and POC: emergency services or emergency room; **or** TTT: emergency and POC: telemedicine | emergency; emergency_ambulance |
| Acute | TTT: within 24 hours and POC: emergency services or emergency room; **or** TTT: immediate or within 24 hours and POC: contract doctor or telemedicine | consultation_24 |
| Not urgent | TTT: not urgent; and POC: contract doctor, telemedicine, or self-care | consultation; self_care |

The Zi consensus provided the preliminary reference label. Records with a non-assignable reference or system recommendation, including missing source values and source combinations outside these rules, were excluded from the respective system-class analysis; exclusions are reported as coverage.

#### Mapping validation

The outcome-mapping rules were developed jointly by JS, LEK and TS and also discussed with various members of the BeST Study Group. Ambiguous recommendations were resolved through discussion with input from the Study Group where required. The rule development and implementation check were conducted without knowledge of the comparative results.

### Statistics and reproducibility

For each triage system and target category, we conducted a one-vs-rest comparison against the Zi label. Sensitivity was the proportion of reference-positive vignettes correctly identified as the target category; specificity was the proportion of reference-negative vignettes correctly not assigned to the target category. We additionally report macro-averaged sensitivity and specificity across the three categories, exact multi-class agreement, coverage, and system-specific confusion matrices. The analysis concept and primary outcome definitions were prespecified in response to the accuracy, reliability, reproducibility, and impartiality requirements under development at the time of study planning, before the results of the consensus process were available; subsequent analyses are identified as sensitivity or exploratory analyses where applicable. Analyses were descriptive; no hypothesis tests, alpha threshold, or P values were used. The unit of analysis was the final physician-redacted vignette. No imputation was performed, and non-evaluable classifications were excluded from the respective system-category analysis and reported as coverage. Performance proportions and confusion-matrix cell percentages reported in the main text use the final frequency weights, Winsorised at the 20th and 80th percentiles and normalised to the known protocol population. Corresponding unweighted performance estimates are provided in Supplementary Tables S1 and S2 for descriptive comparison, with each evaluable vignette contributing equally. These supplementary estimates are reported as point estimates without confidence intervals. Counts of vignettes are reported without weighting. The weighted estimates are interpreted as a regularised representation-weight sensitivity analysis, rather than design-based population estimates.

To quantify resampling uncertainty, we used a vignette-level nonparametric percentile bootstrap, following established guidance for bootstrap confidence intervals in medical statistics^30,31^. For each system, the evaluable vignettes were resampled with replacement and sensitivity, specificity, exact three-class agreement, and macro-averaged sensitivity and specificity were recalculated for each replicate. For weighted analyses, the final Winsorised and normalised vignette weights were retained during resampling and were not re-estimated within bootstrap replicates. Two-sided 95% percentile bootstrap confidence intervals were defined by the 2.5th and 97.5th percentiles of 10,000 replicates. The random-number generator was initialised with a fixed seed. This approach is consistent with bootstrap-based interval estimation for diagnostic accuracy measures^32^, but differs from design-based survey bootstraps that construct replicate weights to reflect a known complex sampling design^33^.

### Reporting guideline and registration

The manuscript was developed and reported with reference to the 2015 Standards for Reporting Diagnostic Accuracy Studies (STARD) and Part B of the Development, Evaluation, and Assessment of Large Language Models (DEAL-B) checklist^34,35^. STARD was interpreted in the context of a vignette-based, multi-class benchmark rather than a conventional clinical diagnostic-accuracy study, and DEAL-B was interpreted in the context of evaluating pretrained models with a controlled prompt and simulated patient interaction. The study was not prospectively registered in a clinical-trial or other public study registry, and no separate study protocol was available for this initial analysis.

## Data availability

The final physician-redacted BeST vignettes will be made publicly available. They contain no direct identifiers or linkable case identifiers. Their vignette-level triage labels will remain undisclosed during the ongoing external expert-consensus process to avoid influencing the participating experts’ assessments. Analyses were run with Python 3.11.16, pandas 3.0.5, NumPy 2.4.6, and Matplotlib 3.11.1. The aggregate results data and analysis code will be published on GitHub before final publication. BeST is open to companies wishing to have their triage systems included in the benchmark; enquiries may be sent to. Information about the benchmark and results from subsequent benchmark rounds will be made publicly available at www.zi.de/BeST. The project page is scheduled to become available with final publication and is not yet online.

## Code availability

The Python analysis code used to generate the reported statistics, tables, and figures will be published on GitHub after publication. English translations of the operational prompts for the LLM workflow are provided in the Supplementary Information as an example illustrating the patient roleplay approach. The scripts used to conduct the role-play interactions are internal implementations and will not be made publicly available. Raw interaction logs will be shared only where permitted by data-governance and licensing requirements.

## Acknowledgments

The authors thank the BeST Study Group for its contributions to the study. The group includes the authors, Daniela Blassfeld, Anne Maciuga, Christopher Pommerenke, Mascha Hochfeld, Karl Schenkel and Thomas Czihal. All authors and named BeST Study Group members are employees of the Central Research Institute of Ambulatory Health Care in Germany (Zi), Berlin, Germany. We declare the use of GPT-5.6 (OpenAI) for language editing of the manuscript.

## Author contributions

Lars Eric Kroll (LEK), Anja Seidel (AS), and Josephine Schuster (JS) contributed equally and share first authorship. AS developed the LLM-based patient role-play workflow for triage benchmarking and the actor-answer confidence-score implementation, and contributed to study design, methodology, software, and data curation. JS led the medical revision and curation of the vignettes and contributed to vignette development, medical validation, and consensus assessment. LEK led the overall study conception, analytical strategy, outcome metrics, formal analysis, software, validation, visualisation, and initial drafting. Edgar Steiger (ES) provided methodological and statistical supervision and contributed to methodology, formal analysis, and validation. Timo Schöpke (TS) provided medical supervision and contributed to medical validation and interpretation. Dominik von Stillfried [DvS] contributed to discussions on the methodological approach, provided relevant literature references, and critically reviewed and revised the manuscript. All authors contributed to critical revision of the manuscript and approved the final version. The BeST Study Group (Daniela Blassfeld [DB], Anne Maciuga [AM], Christopher Pommerenke [CP], Mascha Hochfeld [MH], Karl Schenkel [KS]) contributed together with JS and TS to vignette revision, assessment, and consensus formation. Thomas Czihal [TC] advised on methodological questions and the German Social Code.

## Supplementary information

Supplementary Tables S1 and S2 provide unweighted overall and class-specific performance estimates without confidence intervals. Supplementary Methods present the LLM test workflow and English translations of its German operational prompts as an example illustrating the patient roleplay approach. The remaining system-specific workflows for SmED and Infermedica are internal, unpublished implementations and are not included in the publicly available materials.

