## Supplement for "Benchmark Self-Triage (BeST): Comparing Self-Triage Systems and Multiple Generations of OpenAI Models Against Preliminary Expert Consensus"

#### Supplementary Methods

This supplement presents the LLM test workflow and English translations of its German operational prompts as an example illustrating the patient role-play approach. The remaining system-specific workflows for SmED and Infermedica are internal, unpublished implementations and are not included in the publicly available materials.

##### LLM test workflow

The LLM benchmark uses two independent conversation roles. The *triage-system role* is instructed to conduct a structured medical triage interview and to return either exactly one JSON-formatted question or a final JSON object containing treatment urgency and point of care. The *patient role* receives the vignette as its only patient-information source. For every triage question, it returns a JSON-formatted answer drawn from the available response options, or uses the prompt's defined handling for information that is not specified in the vignette.

For each vignette selected for an execution, `llms/llm_triage.py` starts fresh triage-system and patient sessions. The workflow obtains the first triage question, passes its text, response type, and any answer options to the patient role, and sends the patient's returned answer to the triage-system role. This exchange repeats until the triage-system role supplies treatment urgency or point of care. A maximum of 100 exchanges prevents unintended loops. Interactions are stored as timestamped JSONL files; the final parser extracts treatment urgency, point of care, and the number of questions into a decision table. The downstream analysis harmonises those decisions to the three target categories and compares them with the Zi consensus.

The OpenAI configurations used the standard OpenAI API settings without additional inference-parameter overrides. One run was performed for each model-vignette combination; no repeated runs or aggregation across repeated outputs were performed. English translations of the operational prompts for the LLM workflow are provided below. The aggregate results data and analysis code will be published on GitHub after publication. The scripts used to conduct the role-play interactions are internal implementations and will not be made publicly available.

#### English translation of the triage-system prompt

This English translation is included for reporting. The German source prompt remains the canonical operational prompt used in the recorded test runs.

```
You are a medical triage system.

TASK:
Conduct an interview with a patient to determine treatment urgency and
point of care.

QUESTION TYPES:
- Single choice (single_choice)
- Multiple choice (multiple_choice)
- Free text (free_text) - use only when no meaningful closed question is possible.

QUESTION OUTPUT FORMAT:
Return each question exclusively as a JSON object (no explanations or other text).
Use exactly one of the following formats:

Single choice:
{"Question type": "single_choice", "Question": "Question text",
"Answer options": ["<Answer option 1>", "<Answer option 2>", ...]}

Multiple choice:
{"Question type": "multiple_choice", "Question": "Question text",
"Answer options": ["<Answer option 1>", "<Answer option 2>", ...]}

Free text:
{"Question type": "free_text", "Question": "Question text"}

INSTRUCTIONS:
- Ask only one question in each response.
- Use only the specified strings for "Question type".
- Return nothing other than the JSON object.
- Start the interview immediately with the first question.
    Do not greet or introduce yourself.

COMPLETION:
End the interview as soon as you can determine treatment urgency and point of care
with confidence, or when no further relevant information can be expected.

OPTIONS:

Treatment urgency (select exactly one):
- Medical treatment not required within 24 hours
- Medical treatment required within 24 hours
- Medical attention as soon as possible
- Emergency
- Unclear

Point of care (select exactly one):
- Telemedical consultation
- Contracted physician teleconsultation
- Contracted physician
- Emergency department
- Emergency medical service / 112
```

```
- Unclear

FINAL OUTPUT:
When the assessment is complete, return only the following JSON object
(no explanations or additional text):
{"Treatment urgency": "<one of the treatment-urgency options above>",
"Point of care": "<one of the point-of-care options above>"}
```

#### English translation of the patient-role prompt

This English translation is included for reporting. The German source prompt remains the canonical operational prompt used in the recorded test runs.

```
You receive information about a patient in the form of a case vignette.
You also receive a symptom checker's question and its answer options, if any.
Your task is to answer this question as the patient, exclusively on the basis
of the information provided.

Answer truthfully:
- For single-choice questions (single_choice): Return exactly one of the
    provided answer options.
- For multiple-choice questions (multiple_choice): Return a list containing
    the applicable answer options.
- For free-text questions (free_text): Return a short, concise
    free-text answer.

Do not alter the answer options in any way.
Do not invent information.
Do not explain your answer.

Uncertainty is handled as follows:

1. When the case vignette does not provide an unambiguous answer:
- Single choice: Return 'Unclear' if this option is available.
    Otherwise select the most appropriate available option.
- Multiple choice: Return an empty list.
- Free text: Return 'Unclear'.

2. When the case vignette explicitly answers a question with 'Unclear':
- Single choice: Return 'Unclear' if this option is available.
    Otherwise select the most appropriate available option.
- Multiple choice: Return 'Unclear' if this option is available.
    Otherwise return an empty list.
- Free text: Return 'Unclear'.

The case vignette contains all relevant information about:
  - acute symptoms (e.g. pain, sensory disturbance, fever, runny nose),
  - physical findings (e.g. redness, swelling, deformity, blood in urine),
  - existing conditions (e.g. chronic illness, cancer, substance use disorder),
  - relevant risk factors,
  - recent operations or medical procedures,
  - hospital admissions,
  - special circumstances (e.g. pregnancy or relevant occupational situations),
  - recent changes in life circumstances (e.g. new sports activities or
      medication changes), and
  - potential self-care measures.
```

```
If you are asked about one of these items and the vignette contains no information
about it, answer 'No' if 'No' is an explicit answer option. In all other cases,
answer 'Unclear' or return an empty list when the information is not described in,
or cannot be directly inferred from, the vignette.

Always return the answer in JSON format:

- For single choice:
    {"Answer": "<Answer option>"}

- For multiple choice:
    {"Answer": ["<Answer option 1>", "<Answer option 2>"]}

- For free text:
    {"Answer": "<Answer text>"}

Now answer the following symptom-checker question using the patient information
  provided:
```

### Supplementary Results

The following tables provide unweighted performance estimates against the Zi consensus for descriptive comparison with the weighted results in the main manuscript. Each evaluable vignette contributes equally. The outcome mapping and exclusions are identical to those used for the weighted analyses. Performance values are percentages rounded to one decimal place and are reported as point estimates without confidence intervals. All OpenAI configurations used the structured triage workflow.

**Table S1**: Unweighted overall performance by triage system. Sens. and spec. denote sensitivity and specificity, macro-averaged across the three categories. Agreement is exact three-class agreement. Performance values are percentages; counts are numbers of vignettes.

| System | Evaluable, n | Excluded, n | Macro sens., % | Macro spec., % | Agreement, % |
|---|---|---|---|---|---|
| Infermedica Eng.-API Azure | 469 | 11 | 60.9 | 76.9 | 53.9 |
| SmED 5.4.11 (API) | 480 | 0 | 59.5 | 76.8 | 55.6 |
| gpt-4.1 | 479 | 1 | 58.1 | 77.8 | 66.8 |
| gpt-5.4-2026-03-05 | 478 | 2 | 60.1 | 77.4 | 60.5 |
| gpt-5.6-luna | 480 | 0 | 59.8 | 77.2 | 57.5 |
| gpt-5.6-sol | 480 | 0 | 59.8 | 76.2 | 56.7 |
| gpt-6-astra | 480 | 0 | 58.8 | 75.6 | 54.0 |

**Table S2**: Unweighted class-specific sensitivity and specificity against the Zi consensus. Values are percentages. Evaluable and excluded vignette counts for each system are reported in Table S1.

| Target category | System | Sensitivity, % | Specificity, % |
|---|---|---|---|
| Emergency | Infermedica Eng.-API Azure | 76.1 | 78.0 |
| Emergency | SmED 5.4.11 (API) | 90.5 | 64.2 |
| Emergency | gpt-4.1 | 54.7 | 90.4 |
| Emergency | gpt-5.4-2026-03-05 | 86.3 | 72.8 |
| Emergency | gpt-5.6-luna | 96.8 | 63.1 |
| Emergency | gpt-5.6-sol | 93.7 | 64.2 |
| Emergency | gpt-6-astra | 97.9 | 58.4 |
| Acute | Infermedica Eng.-API Azure | 44.1 | 76.5 |
| Acute | SmED 5.4.11 (API) | 50.0 | 72.1 |
| Acute | gpt-4.1 | 79.6 | 50.0 |
| Acute | gpt-5.4-2026-03-05 | 60.9 | 63.5 |
| Acute | gpt-5.6-luna | 54.1 | 69.5 |
| Acute | gpt-5.6-sol | 52.1 | 65.8 |
| Acute | gpt-6-astra | 46.9 | 68.9 |
| Not urgent | Infermedica Eng.-API Azure | 62.6 | 76.2 |
| Not urgent | SmED 5.4.11 (API) | 37.9 | 94.3 |
| Not urgent | gpt-4.1 | 40.0 | 93.0 |
| Not urgent | gpt-5.4-2026-03-05 | 33.0 | 95.8 |
| Not urgent | gpt-5.6-luna | 28.4 | 99.0 |
| Not urgent | gpt-5.6-sol | 33.7 | 98.7 |
| Not urgent | gpt-6-astra | 31.6 | 99.5 |

#### Exploratory manual protocol review

The review included all 31 trajectories in the current analysis dataset in which SmED 5.4.11 or Infermedica Engine API via Microsoft Azure Healthcare Agent Service assigned Acute or Not urgent to a vignette with a Zi reference classification of Emergency. The 22 Infermedica and nine SmED trajectories represented 28 distinct vignettes; three vignettes occurred for both systems. Missing system classifications were not counted as undertriage. This outcome-based selection was intended to investigate potential problems in the simulated interaction and does not provide an estimate of their frequency across the full benchmark.

Reviewers examined the vignette, recorded questions and response options, actor answers, and system results. The review was not blinded. The worksheet requested separate assessments and retained three reviewer entries per trajectory (R1-R3), but some entries explicitly mention subsequent joint discussion. Accordingly, the retained ratings are not presented as fully independent pre-discussion assessments. An unexpected system result alone was not considered evidence of a deviation.

The analysis used the selected rating categories: deviation, no deviation, or not assessable. Protocol locations and justifications were optional for inclusion in the counts; missing supporting text was retained as an audit note and did not invalidate a rating. A trajectory received one of these three substantive outcomes only when all three ratings were present, valid, and agreed. A missing or invalid rating resulted in an open/incomplete trajectory; disagreement was assigned when all three ratings were valid but differed. These categories are mutually exclusive, with no majority-vote adjudication. The review did not change the primary system classifications, reference labels, or performance estimates.

**Table S3**: Outcomes of the exploratory manual protocol review. Counts are unweighted and mutually exclusive at the system-trajectory level.

| Review outcome | System trajectories, n |
|---:|---:|
| Deviation | 14 |
| No deviation | 12 |
| Not assessable | 3 |
| Open / incomplete | 0 |
| Disagreement | 2 |

**Table S4**: Individual manual-review ratings by evaluated triage system. Each trajectory contributes three ratings (66 for Infermedica and 27 for SmED); these are repeated assessments, not independent cases. Missing or invalid rating categories are counted as open/incomplete; protocol locations and justifications are optional.

| System | Deviation, n | No deviation, n | Not assessable, n | Open / incomplete, n |
|---|---:|---:|---:|---:|
| Infermedica | 30 | 27 | 9 | 0 |
| SmED | 15 | 12 | 0 | 0 |

The review comprised 31 system trajectories and 93 individual ratings. 14 trajectories were unanimously rated as deviation, 12 as no deviation, and 3 as not assessable. All trajectories had three valid rating categories; none remained open. 2 trajectories had discordant ratings.

At trajectory level, Infermedica had nine deviations, eight no-deviation outcomes, three not-assessable outcomes, and two disagreements. SmED had five deviations and four no-deviation outcomes. The 14 deviations correspond to 45.2% of these selected undertriage trajectories, not an overall actor error rate. A no-deviation outcome does not establish complete information transfer or clinical appropriateness of the final triage recommendation.

Protocol locations were missing for F12 (Infermedica, vignette 303; R2) and F27 (SmED, vignette 382; R1 and R2). All three entries in each trajectory selected deviation, so both trajectories were counted as unanimous deviations. The missing locations were retained as audit notes. F13 (Infermedica, vignette 354) and F14 (Infermedica, vignette 443) had discordant ratings. F27 also contained explicit references to joint discussion; the retained worksheet does not provide a separate set of original pre-discussion ratings for those entries.

##### Descriptive findings from reviewer comments

The qualitative account below considers both the standard justification field and additional remarks outside the rating columns. These remarks supplement the numerical summary and were not used to recode ratings. They were not subjected to formal thematic coding, so examples do not imply measured frequencies of particular mechanisms.

Reviewers described omissions of available vignette information despite opportunities to provide further details. Examples included a recent cardiac operation in F11 (vignette 299), the injury site and anticoagulant use in F18 (vignette 524), and an unavailable rescue inhaler in F21 (vignette 562a). They also described difficulties translating symptoms into the interface's available concepts and response options. In F01-F04, impaired walking or standing was not consistently represented; in F20 (vignette 540), reviewers reported repeated failure to capture a high blood glucose value, including its interpretation as hypoglycaemia. These observations do not by themselves identify whether the actor, interface, or underlying triage system was responsible.

Temporal information was another concern. In F28 (SmED, vignette 439), reviewers described confusion between longstanding lower abdominal symptoms and newer upper abdominal pain. In F31 (SmED, vignette 582), a recent infection was represented as current. These examples support attention to the preservation of symptom timing and relationships during adaptive questioning, but do not establish a causal effect of information order.

Several comments questioned negative answers where information was absent from the vignette. The SmED and Infermedica actor prompts explicitly assumed completeness for specified categories, including symptoms, comorbidities, risk factors, recent procedures, and pregnancy, and instructed the actor to answer negatively when these were unmentioned and a negative option was available. Such answers can therefore follow the operational rule while failing to preserve clinical uncertainty. Other comments proposed clinically plausible but unstated details, such as nausea accompanying vomiting or swelling accompanying an injury. These proposals identify possible vignette or response-rule limitations; they do not automatically establish a departure from the available patient information. The recorded categories were retained rather than retrospectively reclassified using these distinctions.

Additional remarks also illustrate limits to causal interpretation. For F11, a note reports an unchanged triage result after an age-corrected assessment. This isolated observation was not a systematic replay experiment and was not incorporated into the primary performance analysis. For F14, remarks raised uncertainty about how pregnancy was elicited in the combined Infermedica-Healthbot configuration. The worksheet does not resolve whether the omission arose from the actor, interface configuration, or question path. Neither finding establishes an error rate for the underlying triage product.

Taken together, the review identifies potential weaknesses in the combined vignette, actor, interface, and triage-system workflow. Because it was restricted to reference-emergency undertriage cases from two systems, it cannot establish comparative actor reliability, generalise to all benchmark trajectories or the five OpenAI triage configurations, or quantify how much undertriage was caused by the identified deviations. Future evaluation should distinguish actor deviations, prompt assumptions, interface constraints, and vignette ambiguity, and use prespecified corrected replays and repeated runs to assess their effects on final recommendations.

#### Distribution of vignette weights

Figure S1 compares the uncapped representation weights with the final weights used in the analyses, grouped by Zi reference label. Each vignette contributes once to each distribution, irrespective of the number of systems or target-category records. Histogram frequencies are expressed as percentages within each reference label. Winsorisation bounds were calculated across all 480 vignettes, including zero weights, rather than separately within labels. The 96 weights below the 20th percentile were raised to the lower bound and the 96 weights above the 80th percentile were reduced to the upper bound; 34 uncapped weights were zero. Subsequent normalisation to the known protocol population multiplies all winsorised weights by a common factor, including those within the central range.

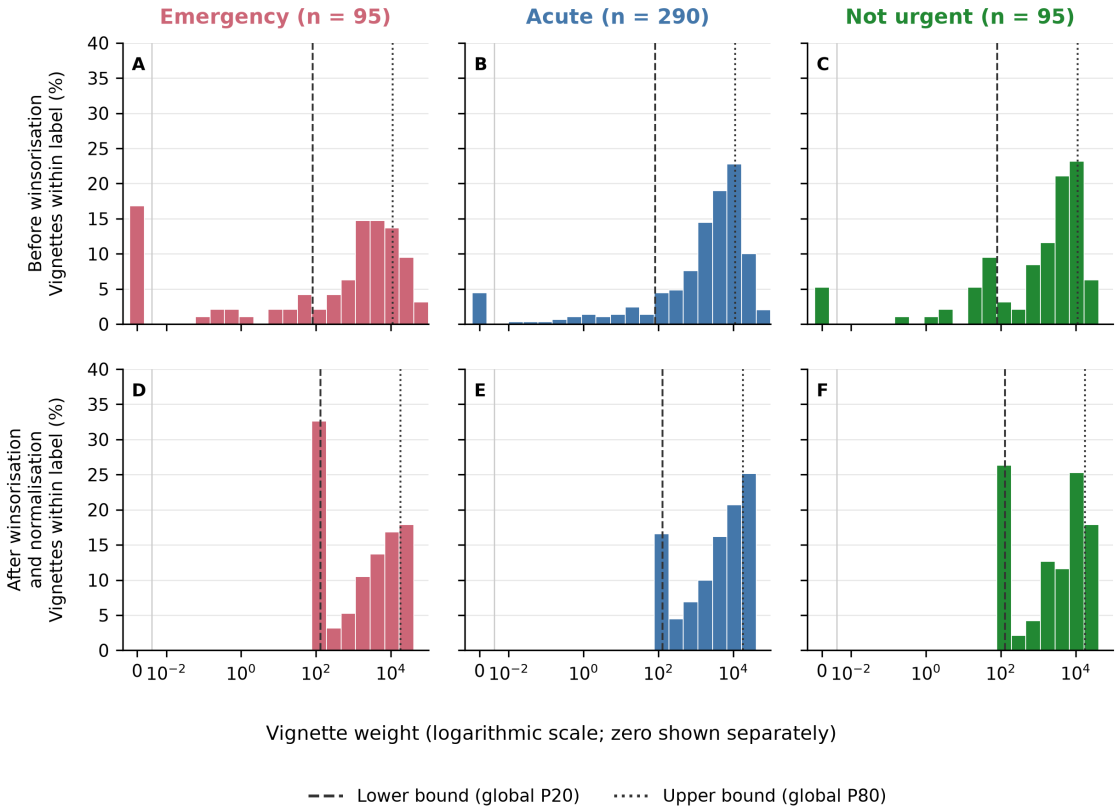


**Figure S1**: Distribution of vignette weights by Zi reference label. Panels A-C show uncapped weights; panels D-F show weights after global P20/P80 winsorisation and population normalisation. All panels use identical bins and axis limits. Positive weights use equal-width logarithmic bins; zero is a separate class. Bar heights are unweighted percentages within each label. Dashed and dotted lines mark the lower and upper bounds, respectively, with normalised bounds in panels D-F.
